# Association between choroidal microvasculature in the eye and Alzheimer’s disease risk in cognitively healthy midlife adults: a pilot study

**DOI:** 10.1101/2024.08.27.24312649

**Authors:** Jamie Burke, Samuel Gibbon, Audrey Low, Charlene Hamid, Megan Reid-Schachter, Graciela Muniz-Terrera, Craig W Ritchie, Baljean Dhillon, John T O’Brien, Stuart King, Ian JC MacCormick, Thomas J MacGillivray

**Affiliations:** Robert O Curle Ophthalmology Suite, Institute for Regeneration and Repair, University of Edinburgh, UK, EH16 4UU; School of Mathematics, University of Edinburgh, Edinburgh, UK, EH9 3FD; Centre for Clinical Brain Sciences, Edinburgh, UK, EH16 4TJ; Department of Psychiatry, School of Clinical Medicine, University of Cambridge, Cambridge, UK, CB2 0SP; Edinburgh Imaging, The Queen’s Medical Research Institute, University of Edinburgh, UK, EH16 4TJ; Heritage College of Osteopathic Medicine, Ohio University, Athens Campus, Athens, Ohio, USA, OH 45701; Princess Alexandra Eye Pavilion, Chalmers Street, Edinburgh, UK, EH3 9HA; Institute for Adaptive and Neural Computation, University of Edinburgh, UK, EH8 9AB

**Keywords:** Optical Coherence Tomography, Retina, Choroid, Dementia, APOE4

## Abstract

**INTRODUCTION:** We explored associations between measurements of the ocular choroid microvasculature and Alzheimer’s disease risk.

**METHODS:** We measured the choroidal vasculature appearing in optical coherence tomography scans of 69 healthy, mid-life individuals in the PREVENT Dementia cohort. The cohort was prospectively split into low, medium, and high-risk groups based on the presence of known risk factors (APOE4 genotype and family history of dementia). We used ordinal logistic regression to test for cross-sectional associations between choroidal measurements and Alzheimer’s disease risk.

**RESULTS:** Choroidal vasculature was progressively larger between ordinal risk groups, and significantly associated with risk group prediction. APOE4 carriers had thicker choroids and larger vascularity compared to non-carriers. Similar trends were observed for those with a family history of dementia.

**DISCUSSIONS:** Our results suggest a potential link between the choroidal vasculature and Alzheimer’s disease risk. However, these exploratory findings should be replicated in a larger sample.

## Introduction

Non-invasive examination of the back of the eye using optical coherence tomography (OCT) is increasingly used to investigate neurodegenerative diseases, including Alzheimer’s disease,^1^ which has a strong vascular component.^2^ While the primary goal of OCT has often been to image the cross-sectional retinal layers (a *neuronal* measure and marker of neurodegeneration), recent advances in enhanced depth imaging OCT (EDI-OCT),^3^ permit visualisation of the choroid, a dense *vascular* mesh posterior to the retina, which provides essential maintenance to the photoreceptors.^4^ In comparison to the retinal circulation, the choroidal circulation has significantly greater blood flow and perfusion pressure,^5,6^ and is innervated by the central autonomic network.^7^ Autonomic dysfunction is common in adults with mild cognitive impairment,^8^ and in those already living with dementia,^9^ which makes the choroid a prime candidate for investigating pathophysiological response in relation to these conditions. Since the advent of EDI-OCT, there have been numerous studies linking choroidal changes to both systemic health outcomes,^10^ and neurological diseases.^11–13^

However, there have been conflicting reports of choroidal changes in established Alzheimer’s disease in older populations,^14–20^ raising questions about the underlying mechanisms and stages of the disease that might influence the choroidal vasculature. One possibility is that genetic factors, such as the apolipoprotein E ε4 (APOE4) allele, and family history of dementia, both of which have been identified as important determinants of Alzheimer’s risk,^21–23^ may play a role. To our knowledge, only one study directly assessed the relationship between choroidal measures and genetic risk (APOE4) in cognitively normal, old-age individuals, but found no evidence of significant differences between carriers and non-carriers.^24^ Choroidal changes in the preclinical or prodromal stage of Alzheimer’s disease remain unexplored, and could provide valuable insights into the underlying mechanisms linking vascular pathology to prospective cognitive decline.

The PREVENT Dementia study,^25^ investigates and tracks concurrent cerebral and retinal vascular changes in a cohort of mid-life individuals, approximately half of whom are at increased risk of Alzheimer’s disease, owing to family history. While most research on ocular microvascular changes in Alzheimer’s disease focuses on older age groups,^14–19,24^ the PREVENT cohort presents a unique opportunity to examine the microvasculature in individuals who may be in the very early preclinical stage of disease.^26^ Accordingly, we conducted an exploratory, pilot study to explore the associations between four choroidal measures (choroidal thickness, total choroidal area, choroidal vascularity index, total vessel area) and two genetic risk factors (APOE4, family history) in cognitively healthy midlife adults at baseline of this study cohort.

## Methods

### Study participants

The protocol for the PREVENT Dementia study has been outlined previously.^25,27^ It aimed to recruit participants aged between 40 and 59 from five sites in the UK and Ireland, and to include a significant portion with a family history of dementia (approximately 50%). Ophthalmic imaging was a sub-study, conducted exclusively at the Edinburgh site, which participants could opt into after cerebrovascular assessment. Participants provided written informed consent and the study adhered to the principles of the Declaration of Helsinki.

Eligibility criteria for the Edinburgh arm was competent and consenting adults recruited from the wider PREVENT study who had the capacity to manoeuvre themselves to the retinal imaging machines unaided and follow instructions to facilitate patient fixation during imaging at baseline. Exclusion criteria were those with a clinical diagnosis of dementia, those without capacity to consent at baseline or individuals not able to fully understand written and verbal English. Exclusion criteria also included participants with current or previous ocular disease affecting the retina or choroid such as glaucoma, macular degeneration, diabetic retinopathy, uveitis, vitreous haemorrhage, pachychoroid, pathological myopia, ischaemic optic neuropathy, optic neuritis or other optic nerve diseases, or those who had undergone previous ocular surgery such as cataract surgery or retinal surgery. These were self-reported at the point of acquisition and any incidental findings were referred to an ophthalmologist for confirmation and subsequent referral.

### Image capture

OCT was captured in both eyes using the Heidelberg spectral domain SPECTRALIS Retina HRA+OCT Module (Heidelberg Engineering, Heidelberg, Germany). Fovea-centred OCT B-scans of the retina and choroid (Supplementary Figure 1A – C) were taken using active eye tracking to prevent artefacts from eye movement, covering a 30° angle at high speed, resulting in approximately 9 mm field of view in the transverse direction. A single, horizontal-line scan was taken with an automatic real time (ART) value of 100 (the number of averaged B-scans taken during a single acquisition) to reduce speckle noise.

Supplementary Figure 1 shows a typical OCT capture with the en face localiser scanning laser ophthalmoscopy (SLO) image in panel (A) and the B-scan in panel (B). B-scans with a signal-to-noise quality index (provided in the HeyEx viewer software, version 1.10.4.0; Heidelberg Engineering, Heidelberg, Germany) of less than 15 were excluded, according to the OSCAR-IB criteria^28^ for retinal OCT quality assessment.

### Choroidal measurements

Measurements of fovea-centred subfoveal choroidal thickness, total choroidal area, total vessel area and choroidal vascularity index (CVI) were computed using *Choroidalyzer*,^29^ a fully automatic deep learning-based image analysis pipeline. Briefly, *Choroidalyzer* automatically segments the choroid region and vasculature of an OCT B-scan (Figure 1A) and detects the fovea (Figure 1B). The choroid was defined as the vascular space between the hyperreflective Bruch’s membrane (anterior) and sclera (posterior). Choroidal thickness was measured as a straight-line micron distance underneath the fovea, locally perpendicular to Bruch’s complex (Figure 1C, blue). Total choroidal area was calculated as the number of pixels within a fovea-centred region of interest (RoI), converted into mm^2^ (Figure 1C, red).

**Figure 1.**
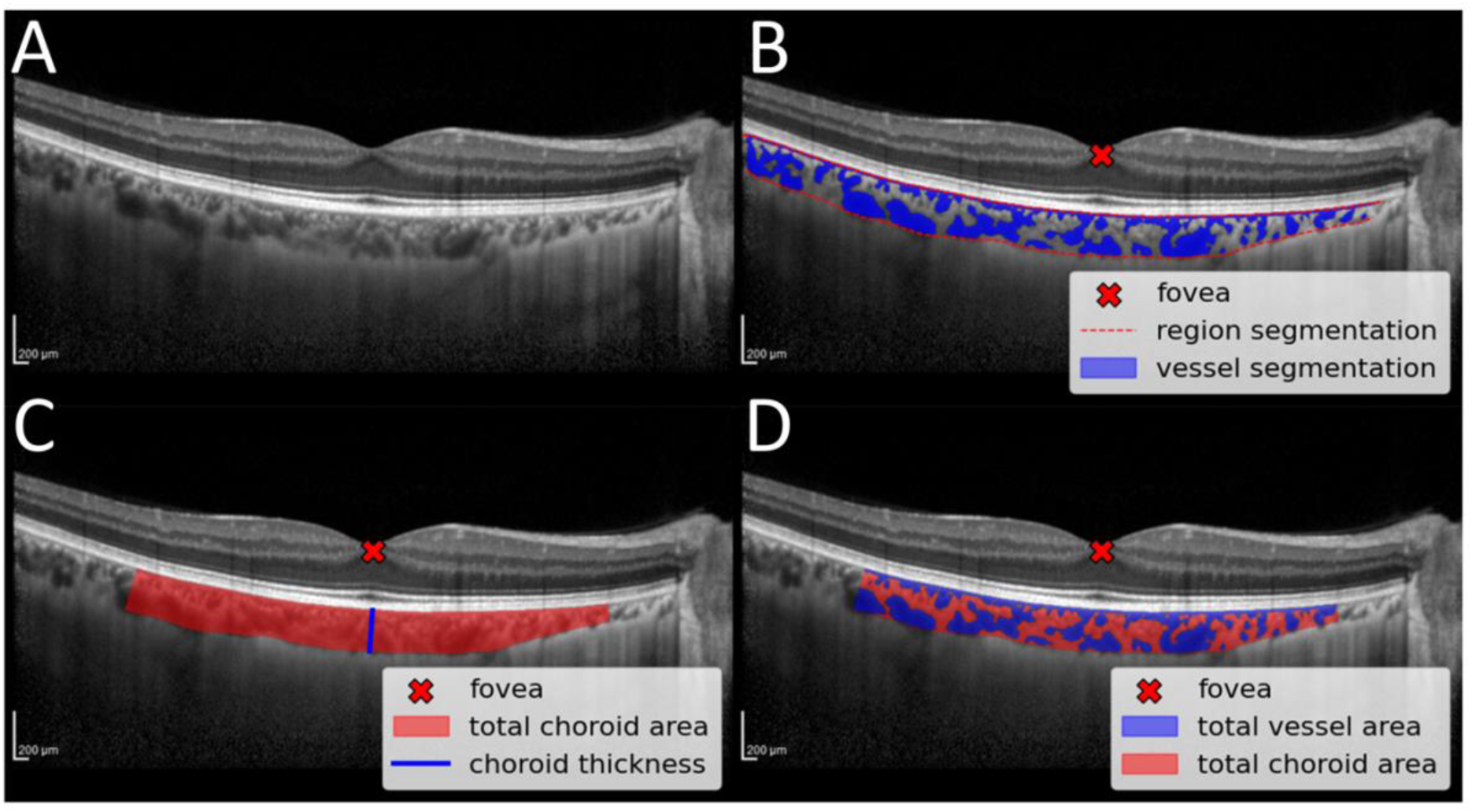
Output from Choroidalyzer and computation of choroidal measurements in a fovea-centred region of interest. (A) OCT B-scan. (B) OCT B-scan with output segmentations from applying Choroidalyzer. (C) Derivation of total choroid area and subfoveal choroid thickness. (D) Derivation of total vessel area and CVI.

Similarly, total vessel area was calculated as the number of vessel pixels within the prescribed RoI (Figure 1D, blue). CVI measures the proportion of vasculature within the choroid and is a dimensionless ratio of vessel pixels to choroid pixels (Figure 1D, blue : red), similarly, measured in a fovea centred RoI. Subsequent region, vessel and fovea detection was checked manually (author J.B.) before measurements were computed. Choroid measurements were made for three distinct RoI’s defined by .5mm, 1.5mm and 3mm distance from the fovea, following the Early Treatment Diabetic Retinopathy Study (ETDRS) grid.^30^ The software is implemented in Python (version 3.11.6).

### Data exploration and statistical analysis

Participants in the PREVENT cohort,^25^ were stratified into three risk groups based on APOE4 presence and family history of dementia (FH) by design: ‘high’ risk was defined where both APOE4 and FH were present, ‘medium’ risk where either APOE4 or FH were present and ‘low’ risk where neither were present.

As this was an exploratory study with a relatively small sample size, our primary objective was to report descriptive statistics and investigate potential trends and associations. We first graphed choroidal measurements, testing for normality using the Shapiro-Wilks test, and performed univariate hypothesis testing on the mean differences between choroidal measures stratified for each risk factor. Alongside univariate statistical tests with individual markers, we assumed a biological ordering between risk groups and used ordinal logistic regression with each choroidal measure in turn as a single covariate in an unadjusted model, as well as in adjusted models controlling for age, sex, and mean arterial blood pressure. We selected these three covariates based on existing literature.^31–34^ Additionally, we included scanning metadata parameters OCT scan quality index (a measure of signal-to-noise ratio) and OCT scan focus (an approximate measure of refractive error) as covariates, to account for differences in image quality and eye shape. Unadjusted models were estimated so as to retain maximum statistical power in a relatively low sample size.

To minimise the risk of spurious associations from analyses of multiple regions of interest per eye, we only selected choroidal measurements from horizontal-line scans with a 1.5mm fovea-centred RoI, with a preference for the left eye. The left eye tends to have a thinner choroid than the right,^35^ helping overcome issues with image quality from optical signal degradation. When the left eye was not available, the right eye was used if available.

Any missing values were removed at the point of analysis (see Table 1). Analyses were performed using Python’s statsmodels (version 0.14.0).

**Table 1.** Demographics and study variables, stratified by risk group.

|  | Risk group |  |  | Overall | P-value |
| --- | --- | --- | --- | --- | --- |
|  | Low | Medium | High |  |  |
| **Participants** | 26 | 26 | 17 | 69 |  |
| **Age** (years) | 50.6 (6.3) | 52.4 (4.5) | 51.6 (6.4) | 51.6 (5.7) | 0.526* |
| **Sex** (female) | 14 (53.9%) | 17 (65.4%) | 10 (58.8%) | 41 (59.4%) | 0.697† |
| **Scan focus** (dioptries) | 0.75 (2.40) | -0.53 (2.52) | -0.68 (1.50) | -0.75 (2.40) | 0.770* |
| **Scan quality** (dB) | 32.5 (3.3) | 33.0 (3.7) | 34.3 (3.1) | 33.2 (3.4) | 0.238* |
| **Hypertension** | 4 (15.4%) | 3 (11.5%) | 1 (6.3%) | 8 (11.8%) | 0.671† |
| **Blood Pressure** (bpm) | 138.6 (13.0) | 124.7 (16.4) | 127.4 (13.7) | 130.6 (15.7) | **0.003\*** |
| **BMI** (kg/m<sup>2</sup>) | 30.4 (6.9) | 28.1 (5.3) | 28.7 (4.2) | 29.1 (5.7) | 0.366* |
| **Smoking** |  |  |  |  | 0.445† |
| Current | 1 (3.9%) | 1 (3.8%) | 0 (0%) | 2 (2.9%) |  |
| Ex | 8 (30.8%) | 7 (26.9%) | 9 (52.9%) | 24 (34.8%) |  |
| Non | 17 (65.4%) | 18 (69.2%) | 8 (47.1%) | 43 (62.3%) |  |
| **Diabetes** | 1 (3.9%) | 2 (7.7%) | 0 (0%) | 3 (4.3%) | 0.475† |
| **Risk factors** |  |  |  |  |  |
| APOE4 status | 0 (0%) | 8 (30.8%) | 17 (100.0%) | 25 (36.2%) | <**0.001**† |
| Family History | 0 (0%) | 18 (69.2%) | 17 (100.0%) | 35 (50.7%) | <**0.001**† |
| **Choroidal measures** |  |  |  |  |  |
| Total Choroid Area (mm<sup>2</sup>) | 0.68 (0.25) | 0.71 (0.194) | 0.87 (0.30) | 0.74 (0.26) | **0.044\*** |
| Choroid Thickness (µm) | 250.39 (84.48) | 257.00 (78.37) | 303.35 (105.56) | 265.93 (89.32) | 0.133* |
| CVI | 0.48 (0.06) | 0.53 (0.07) | 0.53 (0.08) | 0.51 (0.07) | **0.007\*** |
| Total Vessel Area (mm<sup>2</sup>) | 0.31 (0.14) | 0.36 (0.14) | 0.46 (0.18) | 0.37 (0.16) | **0.008\*** |

**Abbreviations:** BMI (body mass index); CVI (choroidal vascularity index). **Notes:** All values are N (%) or Mean (standard deviation). Although modelling below only controlled for age, sex, and blood pressure, we report other cardiovascular risk factors for completeness. Group-based P-values lower than 0.05 are in bold type. \*: One-way ANOVA; †: Chi-squared.
**Missing data (number of eyes, percentage):** Hypertension (1, 1.5%); BMI (2, 2.9%); CVI (2, 2.9%); Vessel area (2, 2.9%).

## Results

The Edinburgh site recruited 224 participants, of which 132 provided written informed consent to the retinal sub-study at baseline. Of these 132 participants, 126 underwent successful OCT capture (device error, N=2; room booking issues, N=4). Participants with retinal pathology observed at the point of acquisition (N=1, macular degeneration) were excluded from our analysis to prevent abnormal retinae from confounding our results, as well as highly myopic/hyperopic eyes (N=4, refractive error > ±6 dioptres, approximately measured using the OCT scan focus on the imaging device),^36^ which have a known association with the choroid.^37^ Only EDI-OCT scans with sufficient choroid visualisation were selected for analysis (N=72). After removing individuals with missing data for APOE4 and FH (N=3), the final sample contained 69 participants (66 left eyes, 3 right eyes), stratified into low (N=26), medium (N=26) and high (N=17) risk groups. A sample derivation flowchart is presented in Figure 2. While we cannot be certain of no selection bias in our final sample, upon comparing any distribution differences between the final sample (N=69) and the remaining cohort (N=155), there was no evidence of significant differences in demographic or cardiovascular variables (Supplementary Table 1).

**Figure 2.**
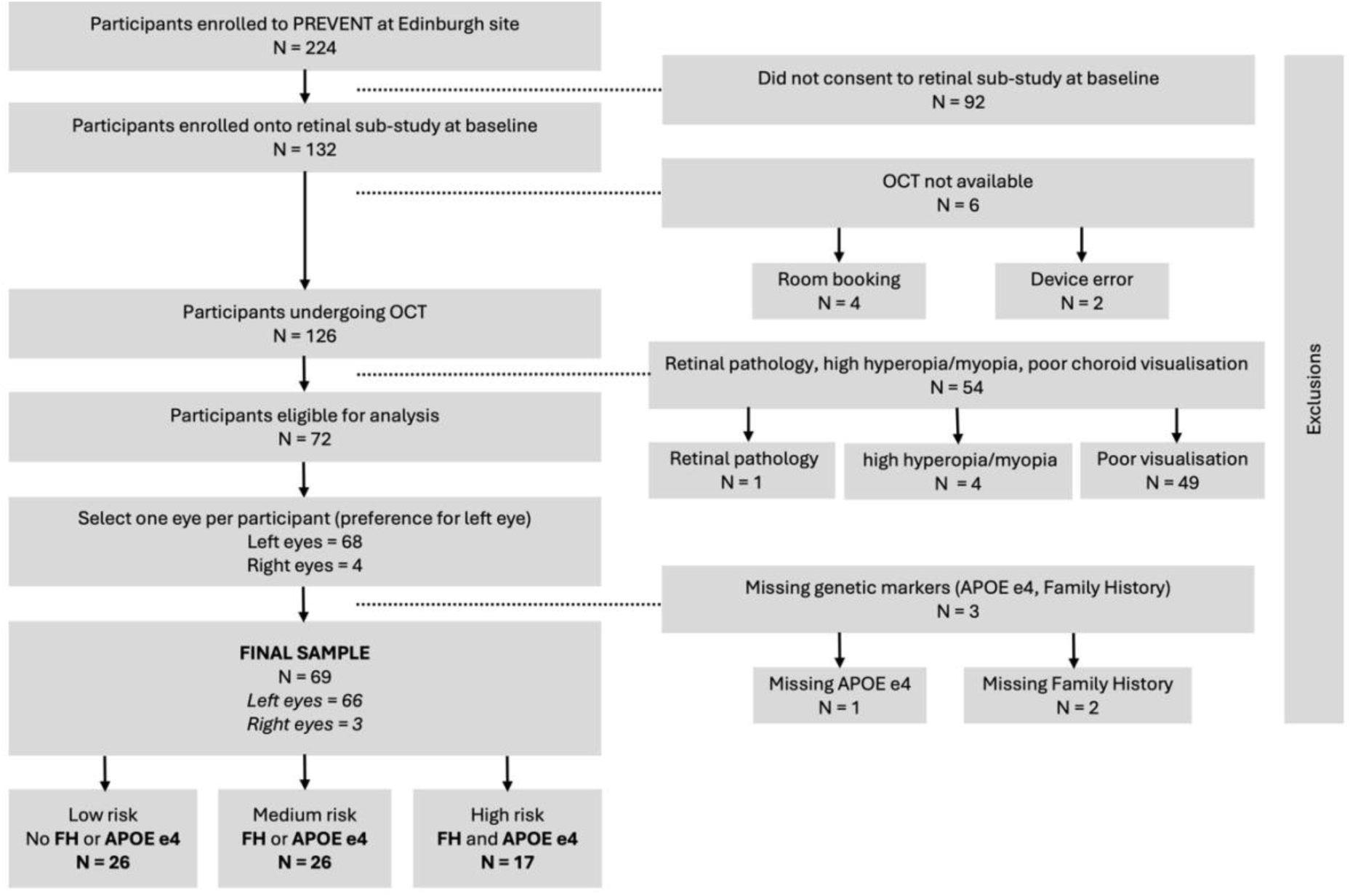
Sample derivation flowchart.

Demographics and study variables are summarised in Table 1. The mean age was 51.6 years (SD = 5.71) with a slight female majority (N = 41, 59.4%). Mean arterial blood pressure was slightly higher in the low-risk group compared to the medium and high-risk groups (p=0.001, p=0.01). No other evidence of significant differences in demographic or cardiovascular variables were observed between risk groups.

Choroidal measurements were normally distributed (Supplementary Figure 2). Figure 3 presents grouped boxplots of choroidal measurements and prospectively defined Alzheimer’s disease risk groups, showing a biological gradient between increasing risk and increasing choroidal measurements, particularly for choroid vessel area.

**Figure 3.**
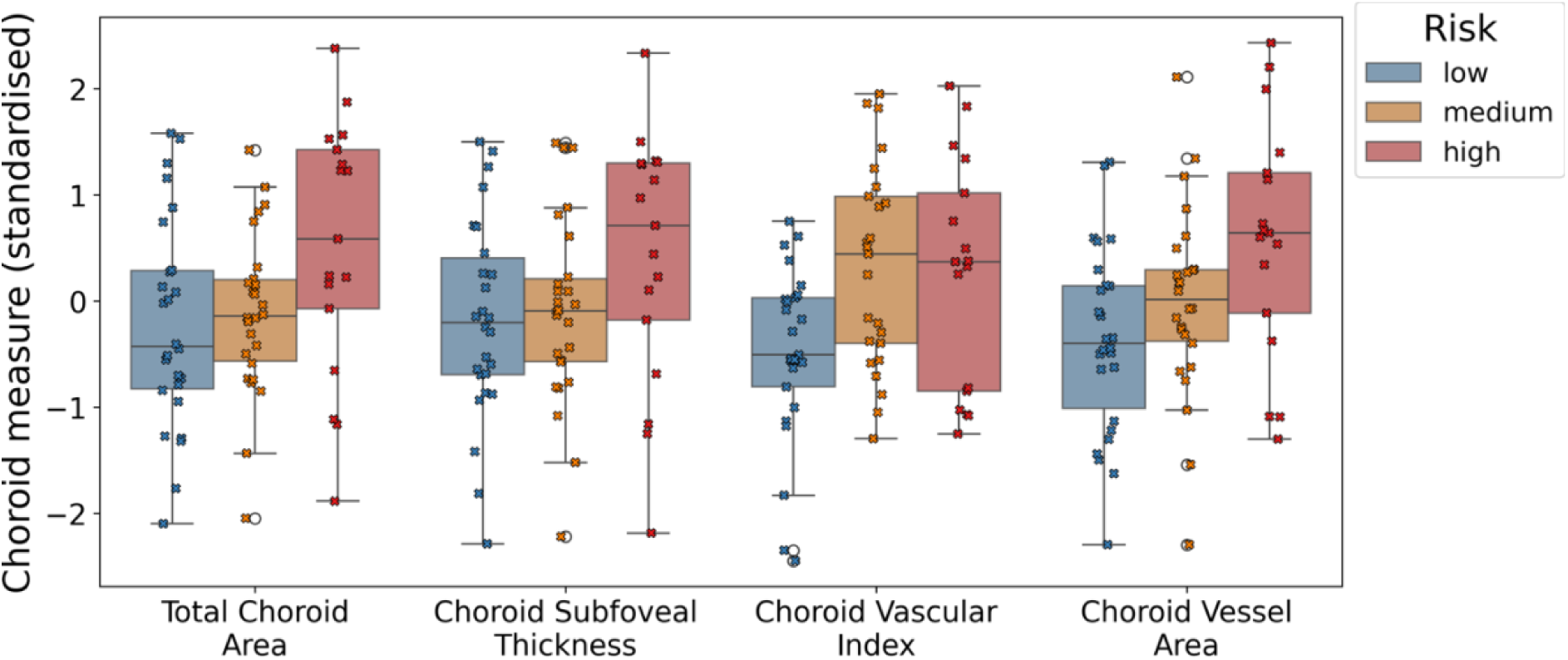
Boxplots showing the relationship between choroidal measures and risk groups, with individual datapoints overlaid in a swarm-plot.

After adjusting for age, sex, blood pressure, OCT scan focus and OCT image quality, larger choroidal measurements were significantly associated with increased risk for Alzheimer’s disease. Specifically , after adjusting for demographic, cardiovascular and scanning parameters, the presence of APOE4 and/or FH was significantly associated with the presence of APOE4 and/or FH was significantly associated with larger total choroidal area (OR per SD increase = 1.84 [CI: 1.08 – 3.16], p = 0.026), and total vessel area (OR per SD increase = 2.28 [CI: 1.30 – 3.99], p = 0.004), but significance was not reached with choroidal thickness (OR per SD increase = 1.63 [0.97 – 2.73], p = 0.064) or CVI (OR per SD increase = 1.63 [0.97 – 2.73], p = 0.065). These findings were also reflected in the unadjusted models.

The magnitude of effect for total vessel area was the largest of all measures. Put another way, for every unit standard deviation increase in total vessel area (0.16 mm^2^), on average it is 2.25 times more likely that this individual has at least one risk marker for Alzheimer’s disease (APOE4, FH) rather than none, after holding all other covariates constant. Results for both adjusted and unadjusted models are summarised in Table 2. Supplementary Figure 3 shows two representative examples from the low-risk group (Supplementary Figure 3A) and high-risk groups (Supplementary Figure 3B), with the choroid in panel (B) presenting with a distinctively larger choroidal space and vascular tissue than the choroid in panel (A) underneath the fovea (red arrows).

**Table 2.** Odds ratios (with 95% confidence intervals and corresponding P-values) for each ordinal logistic regression model, predicting risk group with each choroidal measure in turn.

| Choroid measure | Adjusted models |  | Unadjusted models |  |
| --- | --- | --- | --- | --- |
|  | OR (CI) | P | OR (CI) | P |
| Total Choroid Area (mm<sup>2</sup>) | 1.84 (1.08 – 3.16) | **0.026** | 1.77 (1.09 – 2.88) | **0.021** |
| Choroid Thickness (µm) | 1.63 (0.97 – 2.73) | 0.064 | 1.54 (0.97 – 2.47) | 0.070 |
| CVI | 1.63 (0.97 – 2.73) | 0.065 | 1.87 (1.16 – 3.00) | **0.010** |
| Total Vessel area (mm<sup>2</sup>) | 2.28 (1.30 – 3.99) | **0.004** | 2.14 (1.30 – 3.51) | **0.003** |

**Abbreviations:** CVI (choroidal vascularity index); OR (Odds ratio); CI (95% confidence interval). **Notes:** Adjusted models include age (standardised), blood pressure (standardised), OCT scan focus (standardised), OCT image quality index (standardised) and sex as covariates. To provide stable confidence intervals in logistic models, all choroidal measures were standardised. P-values lower than 0.05 are in bold type.

Considering individual risk, total choroidal area and vascular tissue were larger for APOE4 carriers and FH individuals, and these differences were supported by independent t-tests for total vessel area (p=0.022, p=0.009), and for total choroidal area (p=0.020) and vascular index (p=0.022) for APOE4 carriers and FH, respectively (Table 3). Supplementary Figure 4 illustrates these differences using boxplots and showing distributions.

**Table 3.** Distribution of choroidal measures by dementia risk with P-values for t-tests.

|  | APOE4 carrier status |  | P-Value |
| --- | --- | --- | --- |
|  | Non-carrier | Carrier |  |
| **N** | 47 | 25 |  |
| Total Choroid Area (mm<sup>2</sup>) | 0.67 (0.23) | 0.83 (0.27) | **0.02** |
| Choroid Thickness (µm) | 248.40 (85.36) | 288.64 (94.13) | 0.081 |
| CVI | 0.50 (0.06) | 0.53 (0.08) | 0.104 |
| Total Vessel Area (mm<sup>2</sup>) | 0.33 (0.14) | 0.43 (0.18) | **0.022** |
|  | Family History of Dementia |  | P-Value |
|  | None | Yes |  |
| **N** | 34 | 36 |  |
| Total Choroid Area (mm<sup>2</sup>) | 0.69 (0.23) | 0.79 (0.28) | 0.122 |
| Choroid Thickness (µm) | 252.03 (78.16) | 283.50 (99.83) | 0.146 |
| CVI | 0.49 (0.07) | 0.53 (0.07) | **0.022** |
| Total Vessel Area (mm<sup>2</sup>) | 0.32 (0.15) | 0.42 (0.16) | **0.009** |

**Abbreviations:** CVI (choroidal vascularity index); **Notes:** P-values lower than 0.05 are in bold type.

## Discussion

We explored associations between the choroidal vasculature and two genetic risk factors for Alzheimer’s disease (APOE4, FH) in a mid-life cohort. We observed a significant, positive association between total choroid area and total vessel area with degree of combined risk. To our best knowledge, this is the first exploration of the choroidal vasculature in cognitively healthy midlife adults at increased risk of Alzheimer’s disease. Previous studies have focused only on retinal differences between genetic risk groups for Alzheimer’s disease.^38^

The APOE4 variant is a major risk factor for Alzheimer’s disease,^23^ and has been associated with a wide range of negative health-related outcomes or features including cardiovascular disease,^39^ blood-brain barrier dysfunction,^40^ inflammation,^41^ and autonomic dysfunction.^42^ Lohman, et al.^42^ reports that APOE4 carriers exhibit central autonomic dysfunction in early-stage Alzheimer’s disease, including the parasympathetic control of cardiovascular functions.^43^ Collins et al.^8^ found that mild cognitively impaired patients were on average 5.6 times more likely to have autonomic dysfunction than controls, showing significant parasympathetic deficits, which may be involved in the pathogenesis of hypotension in dementia. Similarly, given that choroidal blood flow is regulated to some extent by autonomic input, particularly parasympathetic-mediated vasodilation, we cautiously hypothesise from our exploratory results that parasympathetic deficits may induce vasodilatory, proinflammatory mechanisms or choroidal hypoperfusion. However, the choroid is a highly heterogeneous vascular compartment, and whether larger vasculature corresponds to larger calibre, or simply more vessels cannot be fully ascertained with current imaging device resolutions.

We found just one other study that used enhanced-depth OCT choroid measures to investigate asymptomatic individuals with known APOE4 status and family history of dementia.^24^ However, in contrast with our findings, Ma et al.,^24^ found no evidence of significant differences in choroidal measures between carriers and non-carriers. One reason for the conflicting findings could be that the Ma study reported choroidal measurements in pixel units. By contrast, we converted choroidal measurements from pixel units into physical units (microns and mm^2^) according to the unique transverse pixel length-scale each B-scan corresponds to, so that measurements across the population could be compared more appropriately. Moreover, the average participant age in the Ma study was around 20 years older than ours, suggesting that age differences may also play a role in the contrasting findings.

Enhanced-depth OCT has previously been used to investigate the link between diagnosed dementia and the choroid, with mixed results. Some have found *smaller* choroidal measurements at the group-level in Alzheimer’s disease and MCI compared to controls.^15–17,20^ Bulut et al.,^17^ and Gharbiya, et al.^16^ showed significant choroidal thinning in Alzheimer’s disease, but selected both eyes per participant for their statistical analyses without accounting for inter-eye correlation at the participant-level, violating a core statistical principle.

Moreover, Gharbiya et al.,^16^ and Bayhan et al.,^15^ analysed smaller cohorts and performed measurement of the choroid manually, which has the potential for measurement error, and may be difficult to reproduce.^44^ Recently, Kwapong et al.,^20^ found lower choriocapillaris density in OCT-Angiography imaging of early-age onset Alzheimer’s disease patients compared with healthy controls, and was able to discriminate between such groups. However, compared to the PREVENT Dementia study, the Kwapong et al,.^20^ cohort was older and already presenting with symptoms sufficient for clinical diagnosis – whether the same approach can be leveraged at the preclinical stage of Alzheimer’s disease still requires further validation.

Other works have observed *larger* choroidal measurements at the group-level in Alzheimer’s disease compared with controls.^18,19^ Asanad et al.,^18^ observed larger choroidal thickness from post mortem human tissue in eight Alzheimer’s disease patients relative to eleven controls using histopathology, while Robbins et al.,^19^ found larger total choroidal area and total vessel area in Alzheimer’s disease compared with controls in enhanced-depth OCT. However, these measurements were reported in pixel space. Moreover, the authors observed significantly lower choroidal thickness in their sample, but these distances were measured without accounting for potential choroidal curvature. Thus, where our results fit into the disease trajectory of Alzheimer’s disease is yet to be determined.

In this exploratory study of asymptomatic people at increased risk of Alzheimer’s disease, we observed trends which, independent of covariates, indicate a larger retinal choroid vasculature in participants who carry the APOE4 genotype or have a family history of dementia, relative to those who do not. This reached statistical significance in choroid vessel area and choroid area. However, subfoveal thickness is less robust in characterizing the choroid than area, and the relationship between vascular index and vessel area/choroid area likely played a role in any significant, distributional differences observed between groups (Supplementary Material: *Interplay of choroidal measurements)*.

Furthermore, measurement error and diurnal variation are unlikely to have had a major effect on our results. The differences in mean choroid measurements between APOE4 carriers and non-carriers (choroidal thickness, 53 microns; total choroidal area, 0.19 mm^2^; CVI, 0.05; and total vessel area, 0.15 mm^2^), were greater than expected from approximate choroidal fluctuation due to diurnal variation, (choroidal thickness, 30 microns; total choroidal area, 0.035 mm^2^; CVI, 0.015; total vessel area, 0.02 mm^2^),^45–49^ the threshold reported on *Choroidalyzer’s* reproducibility for choroidal analysis on single-line OCT B-scans (choroidal thickness, 11.5 microns; total choroidal area, 0.05 mm^2^; CVI, 0.013).^50^ This was also the case for individuals with and without a family history of dementia (Table 3).

The ordinal models give us an indication of the size of the observed associations found in our sample and can be easily interpreted. For example, consider two males of equal age, with the same blood pressure. If the first male’s total vessel area was greater in area by at least 0.16 mm^2^, they are on average over two times more likely to have at least one (or both) of the risk markers, assuming the comparative individual had no genetic markers, thus placing them in a higher risk group for developing later life dementia. Whether this effect exists within the general population should be investigated in cohorts designed to test this hypothesis.

Strengths of the current study include the use of reproducible and fully automatic software, *Choroidalyzer,* to extract measurements from the choroidal vasculature using a standardised region of interest. Further, the software reports values in physical units rather than pixel units, aiding interpretability and helping mitigate spurious associations.^24^ Another strength is the uniqueness of the PREVENT cohort, which permits characterisation of the ocular microvasculature in asymptomatic individuals, at increased genetic risk, and around 20 years younger than most individuals in similar studies.

There were some limitations to this study. Statistical analysis was limited by low power due to small sample size and we cannot be certain that selection bias did not play a role in our results. Lower sample size also prevented the use of mixed models, which can account for the nested structure of eyes within participants. Low sample size also limited the number of covariates. Additionally, participants were primarily white in ethnicity, therefore, generalisability of our findings to other ethnic groups or the wider population may be limited.

Finally, our data collection protocol did not collect measurements of intra-ocular pressure (IOP). As IOP directly affects ocular perfusion pressure (OPP), a key regulator of choroidal blood flow, it could be that the observed changes in choroidal vasculature may be attributed to variations in OPP, acting as a confounding factor independent to Alzheimer’s disease risk. Thus, while our associations could relate to mechanistic links between the choroid microcirculation and the pathogenesis of Alzheimer’s disease, more research is needed with larger sample size and the collection of additional clinically informative variables such as IOP, to better understand and account for potential confounding factors.

However, the PREVENT Dementia study is ongoing, collecting longitudinal data at roughly two-year intervals, allowing future temporal modelling that may further validate our findings.

## Conclusion

In this pilot study, we found that the retinal choroidal vasculature was significantly larger in individuals who were both APOE4 carriers and had a family history of dementia, compared to those with one risk factor or none. These exploratory results suggest a potential association between the retinal choroidal vasculature and Alzheimer’s disease risk, and longitudinal studies are needed to see if the retinal choroidal microvasculature may be predictive of developing future cognitive decline and later life Alzheimer’s disease. Although our findings are exploratory, they provide a foundation for future research that could lead to the development of non-invasive biomarkers for the early detection of Alzheimer’s disease, potentially acting as a primary or secondary endpoint of future clinical trials.

## Acknowledgements

We would like to acknowledge with special thanks to the PREVENT participants, the participant panel, members of the Scientific Advisory Committee, and funders for their support of the PREVENT dementia programme.

J.B. was supported by the Medical Research Council (grant MR/N013166/1) as part of the Doctoral Training Programme in Precision Medicine at the Usher Institute, University of Edinburgh.

S.G. was supported by the UK Biotechnology and Biological Sciences Research Council as part of the EASTBIO Doctoral Training Programme at the University of Edinburgh [grant number: BB/M010996/1].

## Consent statement

All participants provided written informed consent, and the study was carried out in compliance with the Declaration of Helsinki.

## Funding

The PREVENT dementia Programme is funded by the Alzheimer’s Society (grant numbers 178, 264 and 329), Alzheimer’s Association (grant number TriBEKa-17-519007) and philanthropic donations. This work was supported by the Alzheimer’s Drug Discovery Foundation (project no. GDAPB-201808-2016196, “Delivering novel neuro-retinal biomarkers for the early diagnosis of Alzheimer’s disease”); NHS Lothian R&D; and British Heart Foundation Centre for Research Excellence Award III (RE/18/5/34216). The funding sources were not involved in designing, conducting, or submitting this work.

## Declaration

The PREVENT Dementia programme received multi-site ethical approval from the UK London-Camberwell St Giles National Health Service (NHS) Research Ethics Committee (REC reference: 12/LO/1023, IRAS project ID: 88938), which operates according to the Helsinki Declaration of 1975 (and as revised in 1983). All substantial protocol amendments have been reviewed by the same ethics committees and favourable opinion was granted before implementation at sites. The retinal sub study in Edinburgh was approved by the South East Scotland Research Ethics Committee (15/SS/0146)

## Data availability

The PREVENT dataset is available to access through a data request on the study website (www.preventdementia.co.uk); on the Alzheimer’s Disease Data Initiative (ADDI) platform baseline dataset DOI: https://doi.org/10.34688/PREVENTMAIN_BASELINE_700V1; Dementia Platforms UK (DPUK); and the Global Alzheimer’s Association Network (GAAIN).

## Notes

### Competing Interest Statement

J.Burke, S. Gibbon, A. Low, C. Hamid, M. Reid-Schachter, G. Muniz-Terrera, C. W. Ritchie, B. Dhillon, J. T. O'Brien, S. King, I. J. C. MacCormick and T. J. MacGillivray report no disclosures relevant to the manuscript. Author disclosures are available in the supporting information.

### Summary of Updates

Manuscript has been edited according to the peer-review process from journal Alzheimer's & Dementia: Diagnosis, Assessment & Disease Monitoring. These include: - Abstract has been shortened. - Reframing the work in manuscript as a pilot study. - Sample derivation described in more detail. - Including OCT scan focus and OCT scan quality as covariates into the ordinal logistic models. - Including variable intra-ocular pressure not collected as a limitation. - Provided more detail around our hypothesis on the observed choroidal changes related to autonomic dysfunction. - Figure 1 revised to show Choroidalyzer's process more clearly. - Supplemental files updated with more figures including an exemplar OCT scan (Supp. Figure 1), and notable choroidal differences between risk groups (Supp. Figure 3). - Conclusion has been lengthened to highlight potential clinical value of work.

## References

1. Chan VTT, Sun Z, Tang S, et al. Spectral-Domain OCT Measurements in Alzheimer’s Disease: A Systematic Review and Meta-analysis. Ophthalmology. 2019;126(4):497–510. doi:10.1016/j.ophtha.2018.08.009

2. Livingston G, Huntley J, Sommerlad A, et al. Dementia prevention, intervention, and care: 2020 report of the Lancet Commission. The Lancet. 2020;396(10248):413–446. doi:10.1016/S0140-6736(20)30367-6

3. Spaide RF, Koizumi H, Pozonni MC. Enhanced Depth Imaging Spectral-Domain Optical Coherence Tomography. Am J Ophthalmol. 2008;146(4):496–500. doi:10.1016/j.ajo.2008.05.032

4. Nickla DL, Wallman J. THE MULTIFUNCTIONAL CHOROID. Prog Retin Eye Res. 2010;29(2):144–168. doi:10.1016/j.preteyeres.2009.12.002

5. Villa C. Ocular Vascular Changes: Choroidal Thickness as an Early Biomarker for Alzheimer’s Disease? J Pers Med. 2021;11(12):1365. doi:10.3390/jpm11121365

6. Martín ESG, Ramírez JM, Rojas B, et al. Glia and blood retinal barrier: effects of ocular hypertension. In: Cardiovascular Disease II. iConcept Press Ltd; 2014:123–162. Accessed June 19, 2024. https://produccioncientifica.ucm.es/documentos/638e0f432558037fa43f9615?lang=en

7. Reiner A, Fitzgerald MEC, Del Mar N, Li C. Neural control of choroidal blood flow. Prog Retin Eye Res. 2018;64:96–130. doi:10.1016/j.preteyeres.2017.12.001

8. Collins O, Dillon S, Finucane C, Lawlor B, Kenny RA. Parasympathetic autonomic dysfunction is common in mild cognitive impairment. Neurobiol Aging. 2012;33(10):2324–2333. doi:10.1016/j.neurobiolaging.2011.11.017

9. Beishon LC, Hosford P, Gurung D, et al. The role of the autonomic nervous system in cerebral blood flow regulation in dementia: A review. Auton Neurosci. 2022;240:102985. doi:10.1016/j.autneu.2022.102985

10. Steiner M, Esteban-Ortega M del M, Muñoz-Fernández S. Choroidal and retinal thickness in systemic autoimmune and inflammatory diseases: A review. Surv Ophthalmol. 2019;64(6):757–769. doi:10.1016/j.survophthal.2019.04.007

11. Andrade C, Beato J, Monteiro A, et al. Spectral-Domain Optical Coherence Tomography as a Potential Biomarker in Huntington’s Disease. Mov Disord. 2016;31(3):377–383. doi:10.1002/mds.26486

12. Garcia-Martin E, Jarauta L, Pablo LE, et al. Changes in peripapillary choroidal thickness in patients with multiple sclerosis. Acta Ophthalmol (Copenh). 2019;97(1):e77–e83. doi:10.1111/aos.13807

13. Robbins CB, Thompson AC, Bhullar PK, et al. Characterization of Retinal Microvascular and Choroidal Structural Changes in Parkinson Disease. JAMA Ophthalmol. 2021;139(2):182–188. doi:10.1001/jamaophthalmol.2020.5730

14. Cunha JP, Proença R, Dias-Santos A, et al. Choroidal thinning: Alzheimer’s disease and aging. Alzheimers Dement Diagn Assess Dis Monit. 2017;8:11–17. doi:10.1016/j.dadm.2017.03.004

15. Bayhan HA, Aslan Bayhan S, Celikbilek A, Tanık N, Gürdal C. Evaluation of the chorioretinal thickness changes in Alzheimer’s disease using spectral-domain optical coherence tomography. Clin Experiment Ophthalmol. 2015;43(2):145–151. doi:10.1111/ceo.12386

16. Gharbiya M, Trebbastoni A, Parisi F, et al. Choroidal thinning as a new finding in Alzheimer’s disease: evidence from enhanced depth imaging spectral domain optical coherence tomography. J Alzheimers Dis JAD. 2014;40(4):907–917. doi:10.3233/JAD-132039

17. Bulut M, Yaman A, Erol MK, et al. Choroidal Thickness in Patients with Mild Cognitive Impairment and Alzheimer’s Type Dementia. J Ophthalmol. 2016;2016:7291257. doi:10.1155/2016/7291257

18. Asanad S, Ross-Cisneros FN, Barron E, et al. The retinal choroid as an oculovascular biomarker for Alzheimer’s dementia: A histopathological study in severe disease. Alzheimers Dement Diagn Assess Dis Monit. 2019;11:775–783. doi:10.1016/j.dadm.2019.08.005

19. Robbins CB, Grewal DS, Thompson AC, et al. Choroidal Structural Analysis in Alzheimer Disease, Mild Cognitive Impairment, and Cognitively Healthy Controls. Am J Ophthalmol. 2021;223:359–367. doi:10.1016/j.ajo.2020.09.049

20. Kwapong WR, Tang F, Liu P, et al. Choriocapillaris reduction accurately discriminates against early-onset Alzheimer’s disease. Alzheimers Dement. 2024;20(6):4185–4198. doi:10.1002/alz.13871

21. Scarabino D, Gambina G, Broggio E, Pelliccia F, Corbo RM. Influence of family history of dementia in the development and progression of late-onset Alzheimer’s disease. Am J Med Genet Part B Neuropsychiatr Genet Off Publ Int Soc Psychiatr Genet. 2016;171B(2):250-256. doi:10.1002/ajmg.b.32399

22. Mak E, Dounavi ME, Low A, et al. Proximity to dementia onset and multi-modal neuroimaging changes: The prevent-dementia study. NeuroImage. 2021;229(January). doi:10.1016/j.neuroimage.2021.117749

23. Stocker H, Perna L, Weigl K, et al. Prediction of clinical diagnosis of Alzheimer’s disease, vascular, mixed, and all-cause dementia by a polygenic risk score and APOE status in a community-based cohort prospectively followed over 17 years. Mol Psychiatry. 2021;26(10):5812–5822. doi:10.1038/s41380-020-0764-y

24. Ma JP, Robbins CB, Lee JM, et al. Longitudinal analysis of the retina and choroid in cognitively normal individuals at higher genetic risk for Alzheimer disease. Ophthalmol Retina. 2022;6(7):607–619. doi:10.1016/j.oret.2022.03.001

25. Ritchie CW, Ritchie K. The PREVENT study: a prospective cohort study to identify mid-life biomarkers of late-onset Alzheimer’s disease. BMJ Open. 2012;2(6):e001893. doi:10.1136/bmjopen-2012-001893

26. Gibbon S, Low A, Hamid C, et al. Association of optic disc pallor and RNFL thickness with cerebral small vessel disease in the PREVENT-Dementia study. Alzheimers Dement Diagn Assess Dis Monit. 2024;16(3):e12633. doi:10.1002/dad2.12633

27. Ritchie CW, Bridgeman K, Gregory S, et al. The PREVENT dementia programme: baseline demographic, lifestyle, imaging and cognitive data from a midlife cohort study investigating risk factors for dementia. Brain Commun. 2024;6(3):fcae189. doi:10.1093/braincomms/fcae189

28. Tewarie P, Balk L, Costello F, et al. The OSCAR-IB consensus criteria for retinal OCT quality assessment. PloS One. 2012;7(4):e34823. doi:10.1371/journal.pone.0034823

29. Choroidalyzer: An Open-Source, End-to-End Pipeline for Choroidal Analysis in Optical Coherence Tomography | IOVS | ARVO Journals. Accessed June 20, 2024. https://iovs.arvojournals.org/article.aspx?articleid=2793719

30. Early Treatment Diabetic Retinopathy Study Design and Baseline Patient Characteristics: ETDRS Report Number 7. Ophthalmology. 1991;98(5, Supplement):741–756. doi:10.1016/S0161-6420(13)38009-9

31. Margolis R, Spaide RF. A Pilot Study of Enhanced Depth Imaging Optical Coherence Tomography of the Choroid in Normal Eyes. Am J Ophthalmol. 2009;147(5):811–815. doi:10.1016/j.ajo.2008.12.008

32. Li XQ, Larsen M, Munch IC. Subfoveal Choroidal Thickness in Relation to Sex and Axial Length in 93 Danish University Students. Invest Ophthalmol Vis Sci. 2011;52(11):8438–8441. doi:10.1167/iovs.11-8108

33. Polak K, Polska E, Luksch A, et al. Choroidal blood flow and arterial blood pressure. Eye. 2003;17(1):84–88. doi:10.1038/sj.eye.6700246

34. Sansom LT, Suter CA, McKibbin M. The association between systolic blood pressure, ocular perfusion pressure and subfoveal choroidal thickness in normal individuals. Acta Ophthalmol (Copenh). 2016;94(2):e157–158. doi:10.1111/aos.12794

35. Lu J, Zhou H, Shi Y, et al. Interocular asymmetry of choroidal thickness and vascularity index measurements in normal eyes assessed by swept-source optical coherence tomography. Quant Imaging Med Surg. 2022;12(1):781. doi:10.21037/qims-21-813

36. Engineering H. SPECTRALIS Product Family User Manual Sw Ver 7.0. Version 7.; 2022.

37. Liu Y, Wang L, Xu Y, Pang Z, Mu G. The influence of the choroid on the onset and development of myopia: from perspectives of choroidal thickness and blood flow. Acta Ophthalmol (Copenh). 2021;99(7):730–738. doi:10.1111/aos.14773

38. López-Cuenca I, Sánchez-Puebla L, Salobrar-García E, et al. Exploratory Longitudinal Study of Ocular Structural and Visual Functional Changes in Subjects at High Genetic Risk of Developing Alzheimer’s Disease. Biomedicines. 2023;11(7):2024. doi:10.3390/biomedicines11072024

39. Lahoz C, Schaefer EJ, Cupples LA, et al. Apolipoprotein E genotype and cardiovascular disease in the Framingham Heart Study. Atherosclerosis. 2001;154(3):529–537. doi:10.1016/S0021-9150(00)00570-0

40. Montagne A, Nation DA, Sagare AP, et al. APOE4 leads to blood–brain barrier dysfunction predicting cognitive decline. Nature. 2020;581(7806):71–76. doi:10.1038/s41586-020-2247-3

41. Tao Q, Ang TFA, DeCarli C, et al. Association of Chronic Low-grade Inflammation With Risk of Alzheimer Disease in ApoE4 Carriers. JAMA Netw Open. 2018;1(6):e183597. doi:10.1001/jamanetworkopen.2018.3597

42. Lohman T, Kapoor A, Engstrom AC, et al. Central autonomic network dysfunction and plasma Alzheimer’s disease biomarkers in older adults. Alzheimers Res Ther. 2024;16(1):124. doi:10.1186/s13195-024-01486-9

43. Lohman T, Sible I, Kapoor A, et al. Blood Pressure Variability, Central Autonomic Network Dysfunction, and Cerebral Small-Vessel Disease in APOE4 Carriers. J Am Heart Assoc. 2024;13(9):e034116. doi:10.1161/JAHA.123.034116

44. Rahman W, Chen FK, Yeoh J, Patel P, Tufail A, Da Cruz L. Repeatability of Manual Subfoveal Choroidal Thickness Measurements in Healthy Subjects Using the Technique of Enhanced Depth Imaging Optical Coherence Tomography. Invest Ophthalmol Vis Sci. 2011;52(5):2267–2271. doi:10.1167/iovs.10-6024

45. Chakraborty R, Read SA, Collins MJ. Diurnal Variations in Axial Length, Choroidal Thickness, Intraocular Pressure, and Ocular Biometrics. Invest Ophthalmol Vis Sci. 2011;52(8):5121–5129. doi:10.1167/iovs.11-7364

46. Usui S, Ikuno Y, Akiba M, et al. Circadian changes in subfoveal choroidal thickness and the relationship with circulatory factors in healthy subjects. Invest Ophthalmol Vis Sci. 2012;53(4):2300–2307. doi:10.1167/iovs.11-8383

47. Tan CS, Ouyang Y, Ruiz H, Sadda SR. Diurnal Variation of Choroidal Thickness in Normal, Healthy Subjects Measured by Spectral Domain Optical Coherence Tomography. Invest Ophthalmol Vis Sci. 2012;53(1):261–266. doi:10.1167/iovs.11-8782

48. Kinoshita T, Mitamura Y, Shinomiya K, et al. Diurnal variations in luminal and stromal areas of choroid in normal eyes. Br J Ophthalmol. 2017;101(3):360–364. doi:10.1136/bjophthalmol-2016-308594

49. Ostrin LA, Harb E, Nickla DL, et al. IMI—The Dynamic Choroid: New Insights, Challenges, and Potential Significance for Human Myopia. Invest Ophthalmol Vis Sci. 2023;64(6):4. doi:10.1167/iovs.64.6.4

50. Burke J, Engelmann J, Gibbon S, et al. OCTolyzer: Fully automatic analysis toolkit for segmentation and feature extracting in optical coherence tomography (OCT) and scanning laser ophthalmoscopy (SLO) data. Published online July 19, 2024. doi:10.48550/arXiv.2407.14128

