## Supplementary Materials for "Association between choroidal microvasculature in the eye and Alzheimer’s disease risk in cognitively healthy midlife adults: a pilot study"

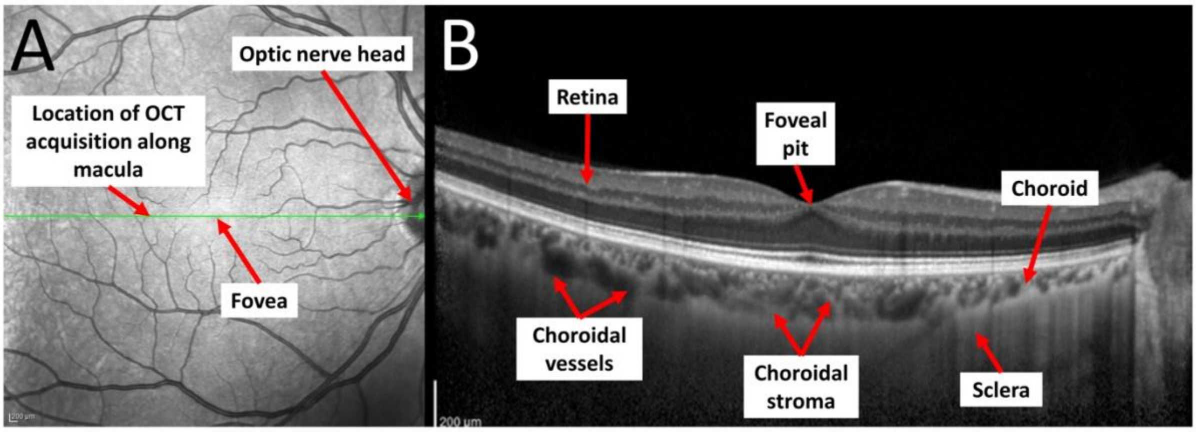

**Supplementary Figure 1.** Exemplar horizontal-line, fovea-centred OCT scan used for analysis with arrows, indicating anatomical landmarks, overlaid. (A) En face localiser SLO. (B) OCT B-scan.

**Supplementary Table 1.** Demographics and study variables of PREVENT Dementia retinal sub-study at the Edinburgh site, stratified by risk group.

| | Overall | Included in final sample | | |
|---|---|---|---|---|
| | | No | Yes | P-Value |
| **Participants** | 224 | 155 | 69 | |
| **Age** (years) | 51.347 (5.532) | 51.255 (5.467) | 51.551 (5.707) | 0.718* |
| **Sex** (female) | 128 (57.658) | 87 (56.863) | 41 (59.420) | 0.834† |
| **Hypertension** | 23 (10.407) | 15 (9.804) | 8 (11.765) | 0.840† |
| **Blood pressure** (bpm) | 128.205 (14.733) | 127.121 (14.204) | 130.594 (15.678) | 0.119* |
| **BMI** (kg/m$^2$) | 28.892 (6.207) | 28.782 (6.415) | 29.143 (5.741) | 0.680* |
| **Smoking** | | | | 0.704† |
| Current | 10 (4.545) | 8 (5.298) | 2 (2.899) | |
| Ex | 78 (35.455) | 54 (35.762) | 24 (34.783) | |
| None | 132 (60.000) | 89 (58.940) | 43 (62.319) | |
| **Diabetes** | 8 (3.620) | 5 (3.289) | 3 (4.348) | 0.707† |
| **Risk factors** | | | | |
| APOE4 status | 89 (40.455) | 64 (42.384) | 25 (36.232) | 0.475† |
| Family History | 104 (47.489) | 69 (46.000) | 35 (50.725) | 0.614† |

**Abbreviations:** BMI (body mass index). **Notes:** All values are N (%) or Mean (standard deviation). **Missing data (number of eyes, percentage):** Sex (2, 0.01%), Age (2, 0.01%), Hypertension (3, 0.01%); Blood Pressure (3, 0.01%), BMI (4, 0.02%); Smoker (4, 0.02%), Diabetes (3, 0.01%), APOE4 (4, 0.02%) and Family History (5, 0.02\%). *, Two-sample t-test; †, Chi-squared test.

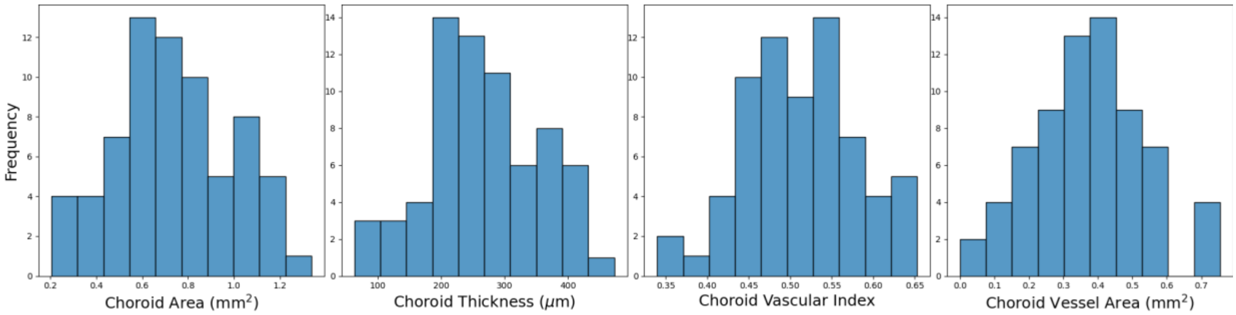

**Supplementary Figure 2.** Histogram distributions of each choroidal measure, approximately normally distributed (Shapiro-Wilks test statistic *t* (P-value) for choroid area: 0.984 (0.5); choroid thickness: 0.979 (0.309); choroid vascular index: 0.984 (0.518); choroid vessel area: 0.987 (0.721)).

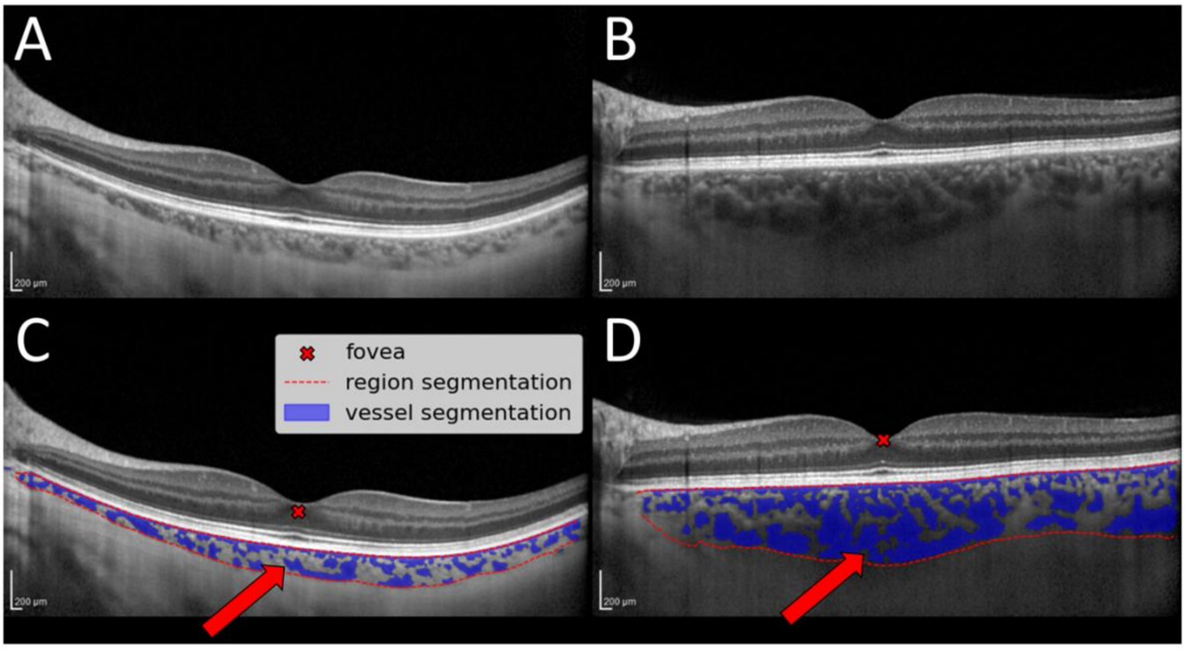

**Supplementary Figure 3**. Exemplar difference in the choroid of a low-risk participant (A) and high-risk participant (B) in terms of total choroid area and total vessel area. Red arrows show a distinct increase in total choroid area and total vessel area underneath the fovea for the high-risk participant relative to the low-risk participant (red cross).

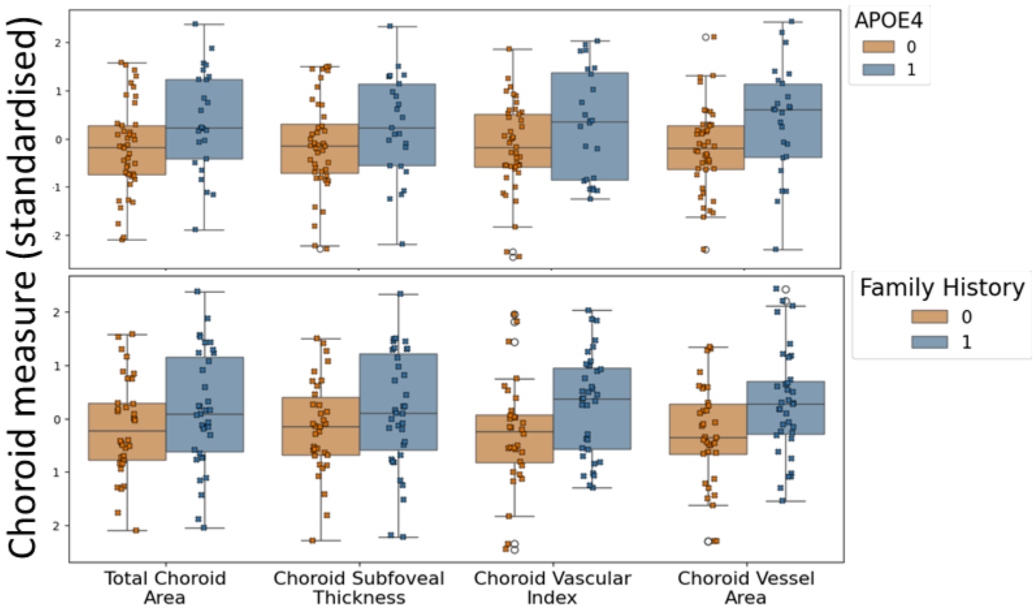

**Supplementary Figure 4.** Boxplots showing the relationship between choroidal measures and APOE e4 carrier status (A), and family history of dementia (B). Both eyes combined

### *Interplay of choroidal measurements*

In this exploratory study, we observed trends which, independent of covariates, indicate an increased choroid vasculature in participants who carry the APOE4 genotype or have a family history of dementia, relative to those who do not. The trends were supported by associations which were statistically significant for all choroidal measures except for (subfoveal) choroidal thickness. Subfoveal choroid thickness is a one-dimensional point-source measurement, which we believe to be unable to characterise subtle choroidal change, particularly in the context of systemic health, relative to two-dimensional measurements such as area and vascular index.

Both choroidal area and vessel area were significantly higher in APOE4 carriers relative to non-carriers. Area and vessel area are directly and inversely proportional to choroidal vascular index (ratio of choroidal vessel area to total choroidal area), respectively, and so these two measures contribute toward estimated associations with choroidal vascular index. Thus, the increase in vascular index (mean increase of 0.03) did not reach significance likely because of the greater mean increase in region area (mean increase of 0.16 $mm^2$) relative to vessel area (mean increase of 0.1 $mm^2$).

For individuals with a family history of dementia, we observed a smaller magnitude of increase in total choroid area relative to APOE4 carriers (mean increase of 0.1 $mm^2$). Put in the context of the significant increase observed in choroidal vessel area (mean increase of 0.1 $mm^2$), this very likely contributed to the significant increase observed for vascular index (mean increase of 0.04). While these observations highlight the interplay between these representative choroidal measurements, they ultimately suggest that choroidal vessel area had the strongest signal among measurements, and that the vasculature itself was the driving factor for increased choroidal area and thickness, rather than surrounding extravascular tissue (for our sample).

*Conflict of Interest*

**Jamie Burke:** None
**Samuel Gibbon**: None
**Audrey Low**: Awarded "Race Against Dementia" Fellowship in past 36 months.
**Charlene Hamid**: None
**Megan Reid-Schachter**: None
**Graciela Muniz-Terrera**: None
**Craig W Ritchie**: Founder, CEO and majority shareholder for "Scottish Brain Sciences". Received consulting fees from "Biogen", "Eisai", "mSD", "Actinogen", "Roche" and "Eli Lilly" in last 36 months. Received payment for presentation-based work by "Roche" and "Eisai" in last 36 months.
**Baljean Dhillon**: None
**John T O'Brien**: Received grant from "Avid/Lilly", "Merck" and "Alliance Medical" in last 36 months. Received consulting fees from "TauRx", "Novo Nordisk", "Biogen", "Roche", "Lilly" and "GE Healthcare" in last 36 months.
**Stuart King**: None
**Ian J.C. MacCormick**: None
**Thomas J. MacGillivray:** None
